# Willingness and influential factors of parents to vaccinate their children against the COVID-19: a systematic review and meta-analysis

**DOI:** 10.1101/2021.08.25.21262586

**Authors:** Petros Galanis, Irene Vraka, Olga Siskou, Olympia Konstantakopoulou, Aglaia Katsiroumpa, Daphne Kaitelidou

**Affiliations:** Clinical Epidemiology Laboratory, Faculty of Nursing, National and Kapodistrian University of Athens, Athens, Greece; Department of Radiology, P & A Kyriakou Children’s Hospital, Athens, Greece; Center for Health Services Management and Evaluation, Faculty of Nursing, National and Kapodistrian University of Athens, Athens, Greece

**Author notes:** **Corresponding author:** Petros Galanis, Assistant Professor, Clinical Epidemiology Laboratory, Faculty of Nursing, National and Kapodistrian University of Athens. **Funding:** None. **Declarations of interest:** None. **Author contributions** P.G and I.V. were responsible for the conception and design of the study. P.G, I.V., O.S., O.K., A.K., and D.K. were responsible for the acquisition, analysis and interpretation of data. All the authors drafted the article or revised it critically for important intellectual content, and provided final approval of the version to be submitted.

**Keywords:** COVID-19, vaccination, willingness, predictors, SARS-CoV-2, children, parents

## Abstract

**Background:** Coronavirus disease 2019 (COVID-19) vaccine uptake among children will be critical in limiting the spread of the severe acute respiratory syndrome coronavirus 2 (SARS-CoV-2) and the disease. Parents are key decision-makers for whether their children will receive a COVID-19 vaccine.

**Objective:** To estimate parents’ willingness to vaccinate their children against the COVID-19, and to investigate the predictors for their decision.

**Methods:** We followed the Preferred Reporting Items for Systematic Reviews and Meta-Analysis guidelines for this systematic review and meta-analysis. We searched Scopus, Web of Science, Medline, PubMed, ProQuest, and CINAHL from inception to August 11, 2021. The review protocol was registered with PROSPERO (CRD42021273125). We applied a random effect model to estimate pooled effects since the heterogeneity was very high. We used subgroup analysis and meta-regression analysis to explore sources of heterogeneity.

**Results:** We found 17 studies including 45,783 parents. The overall proportion of parents that intend to vaccinate their children against the COVID-19 was 56.8% (95% confidence interval: 51.8-61.8%). Parents’ willingness ranged from 29% to 72.7%. Studies quality, sample size, data collection time, and the continent that studies were conducted did not affect the results. The main predictors of parents’ intention to vaccinate their children against COVID-19 were male gender, older age of parents and children, higher socio-economic status, white race, positive attitudes toward vaccination, higher levels of knowledge, and higher levels of perceived threat from the COVID-19, worry, fear, and anxiety.

**Conclusions:** Parents’ willingness to vaccinate their children against the COVID-19 is moderate and several factors affect this decision. Understanding parental COVID-19 vaccine hesitancy does help policy makers to change the stereotypes and establish broad community COVID-19 vaccination. Identification of the factors that affect parents’ willingness to vaccinate their children against COVID-19 will provide opportunities to enhance parents trust in the COVID-19 vaccines and optimize children’s uptake of a COVID-19 vaccine.

## Introduction

Given the human, social and economic burden of the Coronavirus disease 2019 (COVID-19) pandemic, the uptake of a safe and effective vaccine remains a critical strategy to curb its impact (Graham, 2020). Simulation experiments revealed that up to 80% of the population needs to receive a COVID-19 vaccine that is at least 80% effective to largely extinguish the COVID-19 pandemic without any other non-pharmaceutical measures (e.g., social distancing, masks etc.) (Bartsch et al., 2020). Thus, COVID-19 vaccine uptake among children will be instrumental in limiting the spread of the severe acute respiratory syndrome coronavirus 2 (SARS-CoV-2) and the disease.

COVID-19 vaccine uptake relies on adequate production, fair distribution, and high levels of acceptance among the general public (Neumann-Böhme et al., 2020). Recent meta-analyses found that the overall COVID-19 vaccine acceptance rate was approximately 73%, while acceptance among the general population is higher than among healthcare workers (Galanis et al., 2020; Luo et al., 2021; Snehota et al., 2021; Wang, Yang, et al., 2021). Also, real-world data from early studies reveal that COVID-19 vaccination uptake ranges from 28.6% to 98% in the general population (Galanis et al., 2021). Several factors influence vaccination intention and uptake in the general population such as socio-demographic characteristics, attitudes towards vaccination, psychological factors, perceptions of risk and susceptibility to COVID-19, knowledge, information, personal factors, medical conditions, etc. (Al-Amer et al., 2021; Galanis et al., 2021; Snehota et al., 2021; Wake, 2021; Wang, Yang, et al., 2021).

The risk of severe illness and death from the COVID-19 remains quite low for children, but children COVID-19 cases rise sharply due to the highly transmissible delta variant (Tanne, 2021). For instance, since the COVID-19 pandemic began, children represent 14.4% of total COVID-19 cases in the USA but for the week ending August 12, 2021, children were 18% of weekly cases (American Academy of Pediatrics, 2021). Moreover, children make up about 2.4% of total hospitalizations in the USA and about 1% of all pediatric COVID-19 cases resulted in hospitalization since the start of the pandemic (American Academy of Pediatrics, 2021). Thus, there is a need for safe and effective COVID-19 vaccines for children of all ages as swiftly as possible (Tanne, 2021).

Currently, COVID-19 vaccines are approved for children aged 12 and older and it is anticipated that younger children will become eligible since pharmaceutical companies are running clinical trials with children to study the safety and efficacy of COVID-19 vaccines (European Medicines Agency, 2021a, 2021b; Health Canada, 2021).

Since parents are key decision-makers for whether their children will receive a COVID-19 vaccine, it is important to measure willingness of parents to vaccinate their children against the COVID-19. Early studies have already investigated parents’ intention to vaccinate their children but until now, no systematic review on this field is published. Thus, we performed a systematic review and meta-analysis to estimate parents’ willingness to vaccinate their children against the COVID-19, and to investigate the predictors for their decision.

## Methods

### Data sources and strategy

We performed a systematic review and meta-analysis, applying the Preferred Reporting Items for Systematic Reviews and Meta-Analysis (PRISMA) guidelines (Moher et al., 2009). We searched Scopus, Web of Science, Medline, PubMed, ProQuest, and CINAHL from inception to August 11, 2021. We used the following strategy in all fields: ((vaccin*) AND (COVID-19)) AND (parent*). The review protocol was registered with PROSPERO (CRD42021273125).

### Selection and eligibility criteria

Firstly, we removed duplicates, and then we screened consecutively titles, abstracts, and full texts. Also, we examined reference lists of all relevant articles. Two independent researchers performed study selection and a third, senior researcher resolved the discrepancies. We included quantitative studies reporting parents’ willingness to vaccinate their children against the COVID-19. Also, we included quantitative studies that examine factors that affect parents’ willingness to vaccinate their children. Study population included parents and guardians of children aged <18 years. We did not apply criteria regarding study population, e.g. gender, age, race, sample size, etc. Studies published in English in journals with peer review system were eligible to be included. We excluded protocols, reviews, case reports, editorials, and letters to the Editor.

### Data extraction and quality assessment

Two authors independently extracted the following data from the studies: reference, country, data collection time, sample size, age of parents and children, population, study design, sampling method, response rate, percentage of parents that agree to vaccinate their children, and factors that affect parents’ willingness to vaccinate their children. Studies quality was assessed with the Joanna Briggs Institute critical appraisal tool (Santos et al., 2018).

### Statistical Analysis

Parents’ intention to vaccinate their children was assessed with statements or questions like these “When a vaccine for Coronavirus becomes available, I will have my child get it”, “If a COVID-19 vaccine are safe and available to your child for free, how likely would your child be to get vaccinated?”, “At this moment, are you willing to receive COVID-19 vaccination for your child?” etc. Possible answers were in Likert scales (e.g. strongly disagree; disagree; neither disagree nor agree; agree; strongly agree) or in yes/no/uncertain options. For each study, we followed the authors’ decision regarding the positive answer of parents. For instance, in studies where authors used Likert scales, a positive answer could be only one answer (strongly agree) or two answers (agree and strongly agree). We divided the positive answers of parents with the total number of parents to calculate the proportion of parents that agreed to vaccinate their children. Then, we transformed the proportions with the Freeman-Tukey Double Arcsine method and we calculated the proportion of parents that intend to vaccinate their children against the COVID-19 and the 95% confidence interval (CI) (Barendregt et al., 2013).

We used the Hedges Q statistics and I^2^ to assess heterogeneity between studies. A p-value<0.1 for the Hedges Q statistic indicates statistically significant heterogeneity, while I^2^ value higher than 75% indicates high heterogeneity (Higgins, 2003). We applied a random effect model to estimate pooled effects since the heterogeneity between results was very high (Higgins, 2003). We considered sample size, data collection time, age of parents and children, sampling method, response rate, studies quality, and the continent that studies were conducted as pre-specified sources of heterogeneity. Due to the limited number of studies, we decided to perform subgroup analysis for studies quality and the continent that studies were conducted. Also, we performed meta-regression analysis using sample size and data collection time as the independent variables. We treated data collection time as a continuous variable giving the number 1 for studies that were conducted in January 2020, the number 2 for studies that were conducted in February 2020 etc. We conducted a leave-one-out sensitivity analysis to determine the influence of each study on the overall effect. We used the funnel plot and the Egger’s test to assess the publication bias. Regarding the Egger’s test, a P-value<0.05 indicating publication bias (Egger et al., 1997). We did not perform meta-analysis for the factors that influence parents’ decision to vaccinate their children against the COVID-19 since the data were highly heterogeneous and limited. We used OpenMeta[Analyst] for the meta-analysis (Wallace et al., 2009).

## Results

### Identification and selection of studies

After initial search, we found 4325 unique records. Applying the inclusion and exclusion criteria, we identified 17 articles (Figure 1).

**Figure 1.**
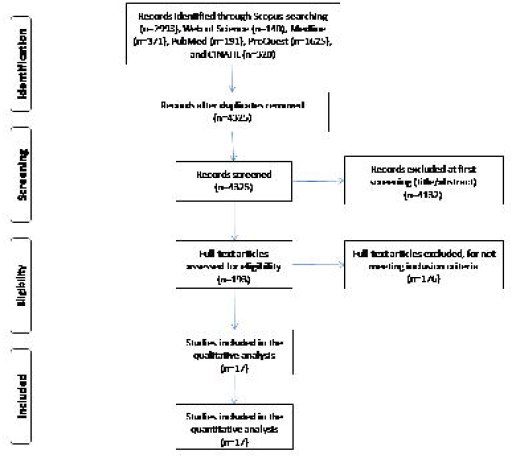
Flowchart of the literature search according to the Preferred Reporting Items for Systematic Reviews and Meta-Analysis.

### Characteristics of the studies

We found 17 studies including 45,783 parents. Details of the studies included in this systematic review are presented in Table 1. Five studies were conducted in the USA (Kelly et al., 2021; Ruggiero et al., 2021; A. M. Scherer et al., 2021; Szilagyi et al., 2021; Teasdale et al., 2021), three studies in China (Wang, Xiu, et al., 2021; Y. Xu et al., 2021; Zhang et al., 2020), two studies in Turkey (Yigit et al., 2021; Yilmaz & Sahin, 2021), one study in Canada (Hetherington et al., 2021), one study in New Zealand (Jeffs et al., 2021), and three studies in Europe (United Kingdom, Germany and Italy) (S. Bell et al., 2020; Brandstetter et al., 2021; Montalti et al., 2021). Also, two studies were multicenter including participants from several countries (Goldman et al., 2020; Skjefte et al., 2021). Data collection time among studies ranged from March 2020 (Goldman et al., 2020) to April 2021 (A. M. Scherer et al., 2021). Sample size ranged from 427 (Ruggiero et al., 2021) to 17,054 parents (Skjefte et al., 2021). All studies were cross-sectional, while 14 studies used a convenience sample (S. Bell et al., 2020; Brandstetter et al., 2021; Goldman et al., 2020; Hetherington et al., 2021; Jeffs et al., 2021; Montalti et al., 2021; A. M. Scherer et al., 2021; Skjefte et al., 2021; Teasdale et al., 2021; Wang, Xiu, et al., 2021; Y. Xu et al., 2021; Yigit et al., 2021; Yilmaz & Sahin, 2021; Zhang et al., 2020), two studies used a probability sample (Kelly et al., 2021; Szilagyi et al., 2021), and one study used the snowball sampling method (Ruggiero et al., 2021).

**Table 1.**
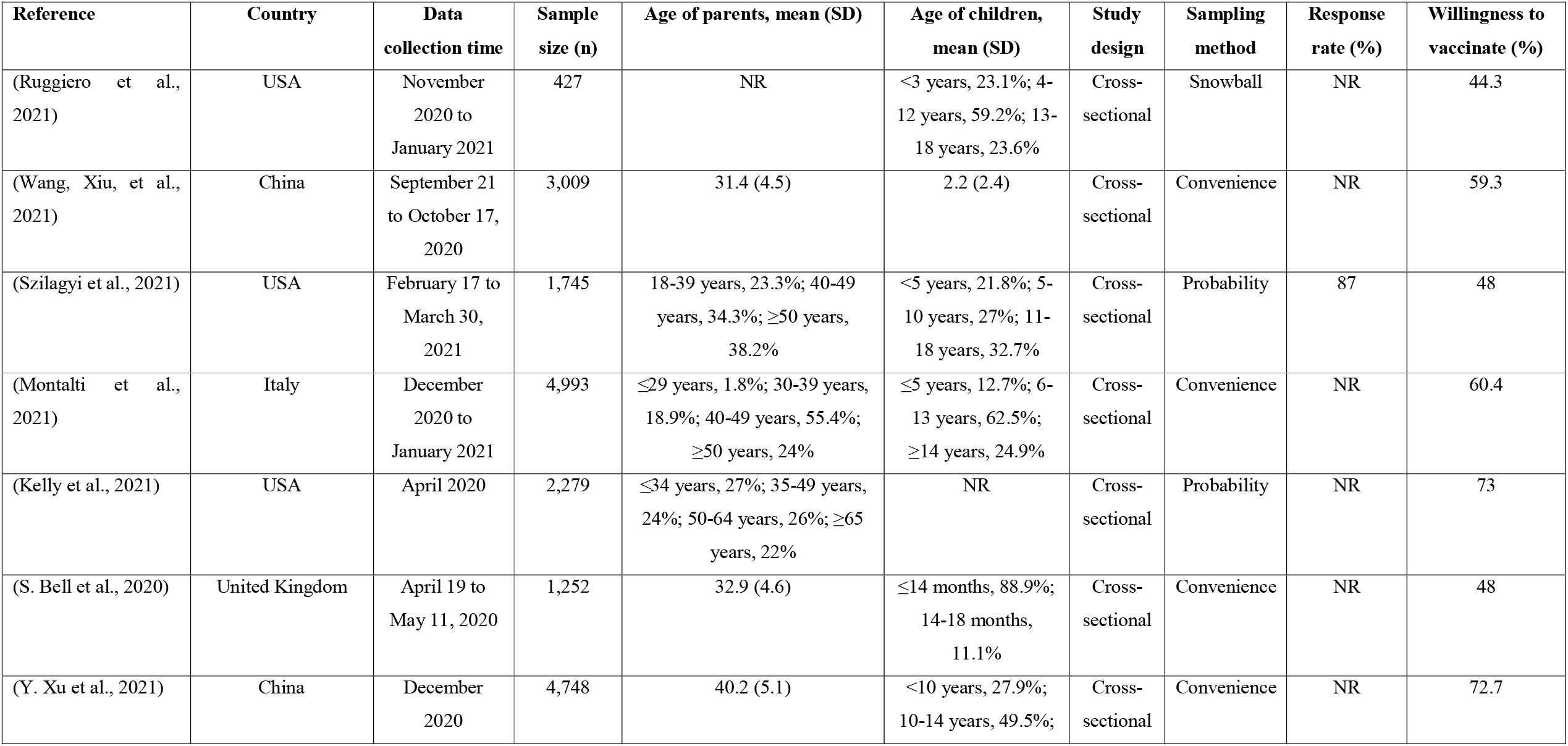
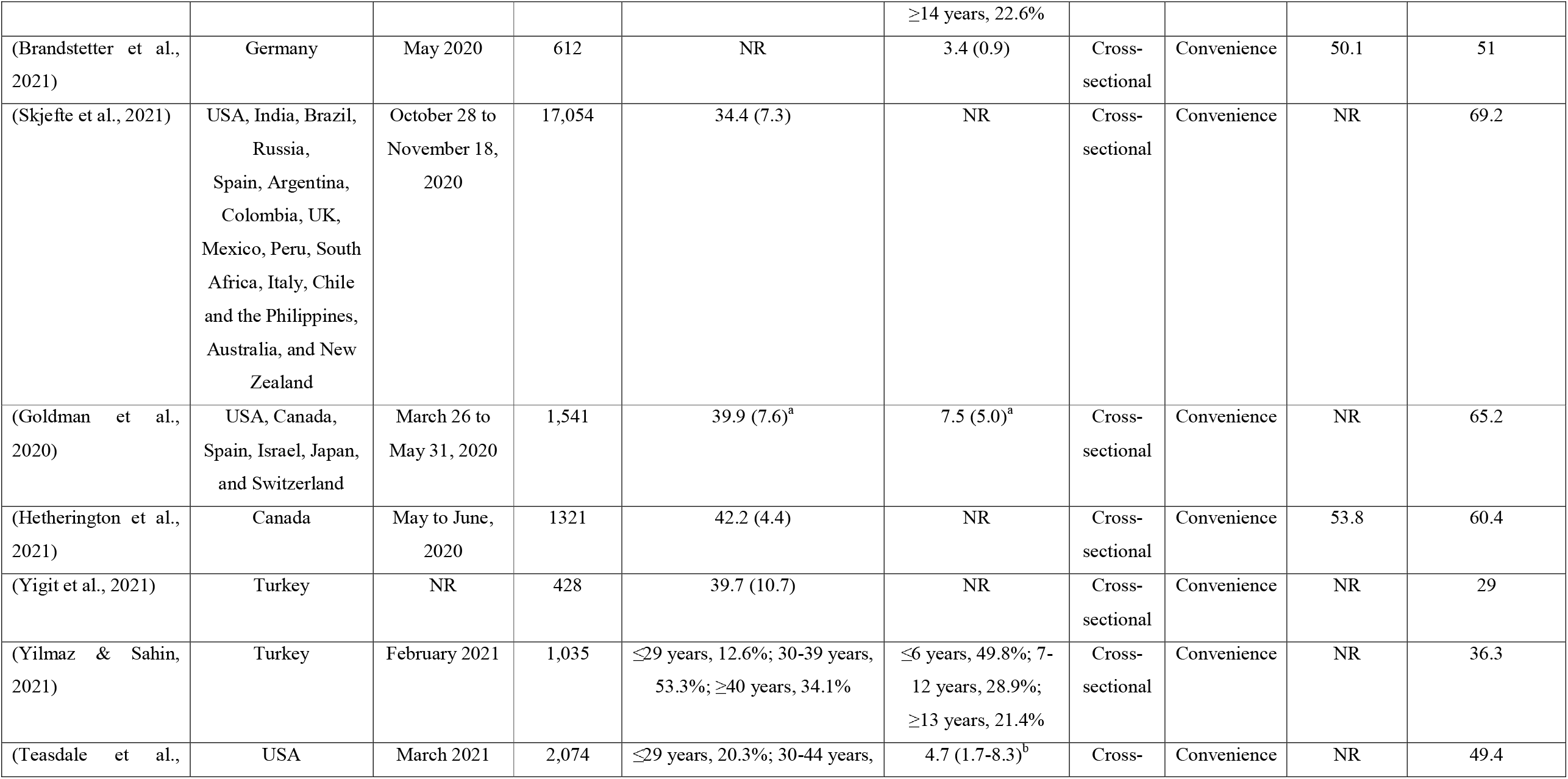
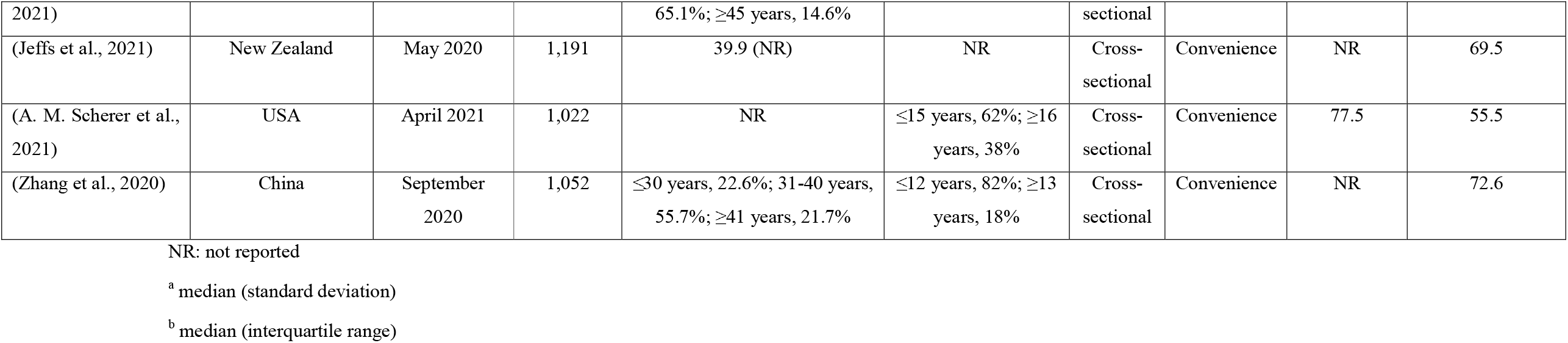
Overview of the 17 studies included in this systematic review.

Thirteen studies did not report data regarding response rate (S. Bell et al., 2020; Goldman et al., 2020; Jeffs et al., 2021; Kelly et al., 2021; Montalti et al., 2021; Ruggiero et al., 2021; Skjefte et al., 2021; Teasdale et al., 2021; Wang, Xiu, et al., 2021; Y. Xu et al., 2021, 2021; Yigit et al., 2021; Yilmaz & Sahin, 2021; Zhang et al., 2020), five regarding children’s age (Hetherington et al., 2021; Jeffs et al., 2021; Kelly et al., 2021; Skjefte et al., 2021; Yigit et al., 2021), and three regarding parents’ age (Brandstetter et al., 2021; Ruggiero et al., 2021; A. M. Scherer et al., 2021).

### Quality assessment

Quality assessment of cross-sectional studies included in this review is shown in Table 3. Quality was good in 14 studies (S. Bell et al., 2020; Brandstetter et al., 2021; Goldman et al., 2020; Kelly et al., 2021; Montalti et al., 2021; Ruggiero et al., 2021; A. M. Scherer et al., 2021; Skjefte et al., 2021; Szilagyi et al., 2021; Teasdale et al., 2021; Wang, Xiu, et al., 2021; Y. Xu et al., 2021; Yilmaz & Sahin, 2021), and moderate in three studies (Hetherington et al., 2021; Jeffs et al., 2021; Yigit et al., 2021). Quality assessment of studies is shown in Supplementary Table 1.

### Meta-analysis

The overall proportion of parents that intend to vaccinate their children against the COVID-19 was 56.8% (95% CI: 51.8-61.8%) (Figure 2). The heterogeneity between results was very high (I^2^=99.08%, p-value for the Hedges Q statistic<0.001). Parents’ willingness ranged from 29% (Yigit et al., 2021) to 72.7% (Y. Xu et al., 2021). A leave-one-out sensitivity analysis showed that no single study had a disproportional effect on the overall proportion, which varied between 55.8% (95% CI: 50.6-61.0%), with Kelly et al. (2021) excluded, and 58.5% (95% CI: 53.7-63.3%), with Yigit et al. (2021) excluded (Supplementary Figure S1). P-value for Egger’s test (<0.05) and funnel plot (Supplementary Figure S2) indicated potential publication bias.

**Figure 2.**
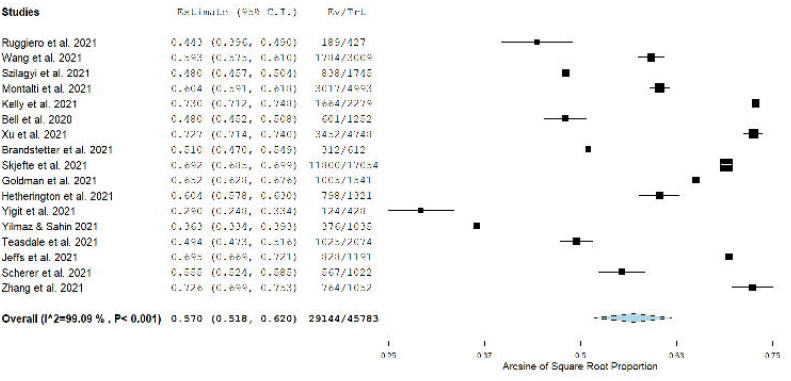
Forest plot of parents’ willingness to vaccinate their children against the COVID-19.

According to subgroup analysis, there were no differences regarding studies quality and the continent that studies were conducted. In particular, the proportion of parents that intend to vaccinate their children in studies that were conducted in North America was 55.2% (95% CI: 45.6-64.7%, I^2^=98.8), in Asia was 54.1% (95% CI=39.8%-68.3%, I^2^=99.5%), and in Europe was 53.2% (95% CI=44.6%-62.2%, I^2^=97.3%). Moreover, the proportion was almost the same for the studies with good quality (57.6% [95% CI: 52.3-62.9%], I^2^=99.16) and those with moderate quality (53.0% [95% CI: 33.2-72.8%], I^2^=99.2). Meta-regression analysis showed that parents’ intention to vaccinate their children was independent sample size (p=0.087) and data collection time (p=0.083).

### Factors related with parents’ willingness to vaccinate their children against COVID-19

Fifteen studies investigated factors that affect positively parents’ willingness to vaccinate their children against COVID-19, while seven studies investigated factors with a negative impact. Statistically significant factors related with parents’ intention to vaccinate their children against the COVID-19 are shown in Table 2. Two studies used univariate analysis (Goldman et al., 2020; Yigit et al., 2021), while the other 15 studies used multivariate analysis eliminating confounders.

**Table 2.** Statistically significant factors related with parents’ willingness to vaccinate their children against the COVID-19.

| Reference | Positive factors | Negative factors | Level of analysis |
| --- | --- | --- | --- |
| (Ruggiero et al., 2021) | Recent history of children's vaccination against influenza, confidence in vaccines, children with chronic illness | Overall vaccination hesitancy, concerns for serious side effects and effectiveness | Multivariate |
| (Wang, Xiu, et al., 2021) | Lower educational level |  | Multivariate |
| (Szilagyi et al., 2021) | Older children, higher educational level, COVID-19 vaccine uptake |  | Multivariate |
| (Montalti et al., 2021) | Fathers, higher educational level, older age | Information based in the web/social media, overall vaccination hesitancy | Multivariate |
| (Kelly et al., 2021) | Fathers, older age, higher educational level, higher income, recent history of parents' vaccination against influenza, increased worry about getting infected, increased perceived threat from the COVID-19 | Black parents | Multivariate |
| (S. Bell et al., 2020) | Increased number of children | Parents from Black, Asian or minority ethnic groups | Multivariate |
| (Y. Xu et al., 2021) |  | Psychological distress | Multivariate |
| (Brandstetter et al., 2021) | Higher educational level, knowledge about prevention measures, high level of information about the COVID-19 pandemic | Stronger beliefs that policy measures were exaggerated, consideration of family or friends as risk group members | Multivariate |
| (Skjefte et al., 2021) | Older age, higher income, higher educational level, health insurance, confidence in COVID-19 vaccine safety and efficacy, belief in the importance of vaccines/mass vaccination, children's complete vaccination history, increased perceived threat from the COVID-19, trust of public health agencies/health science, compliance to mask guidelines |  | Multivariate |
| (Goldman et al., 2020) | Fathers, older children, children with no chronic illness, children's complete vaccination history, recent history of parents' vaccination against influenza |  | Univariate |
| (Hetherington et al., 2021) | Higher educational level, higher income, children's complete vaccination history |  | Multivariate |
| (Yigit et al., 2021) | Fathers, lower educational level, higher levels of fear and anxiety |  | Univariate |
| (Yilmaz & Sahin, 2021) | Healthcare workers, parents' willingness to receive a COVID-19 vaccine, increased perceived threat from the COVID-19, confidence in COVID-19 vaccine safety and efficacy |  | Multivariate |
| (Teasdale et al., 2021) | Fathers, higher educational level, higher income, Asian parents |  | Multivariate |
| (A. M. Scherer et al., 2021) | Fathers, higher educational level | Black parents | Multivariate |
| (Zhang et al., 2020) | Confidence in COVID-19 vaccine safety and efficacy, higher exposure to positive information related to COVID-19 vaccination | Higher exposure to negative information related to COVID-19 vaccination | Multivariate |

Several socio-demographic characteristics affected parents’ intention to vaccinate their children against COVID-19. Higher socio-economic status was related with an increase in COVID-19 vaccine acceptance. In particular, higher educational level (Brandstetter et al., 2021; Hetherington et al., 2021; Kelly et al., 2021; Montalti et al., 2021; A. M. Scherer et al., 2021; Skjefte et al., 2021; Szilagyi et al., 2021; Teasdale et al., 2021), higher income (Hetherington et al., 2021; Kelly et al., 2021; Skjefte et al., 2021; Teasdale et al., 2021), and health insurance (Skjefte et al., 2021) were associated with parents’ intention to accept COVID-19 vaccination for their children. Increased intended uptake of a COVID-19 vaccine was associated with older age of children (Goldman et al., 2020; Szilagyi et al., 2021) and parents (Kelly et al., 2021; Montalti et al., 2021; Skjefte et al., 2021), and increased number of children (S. Bell et al., 2020). Intention of parents to vaccinate their children against COVID-19 increased, when fathers completed the survey (Goldman et al., 2020; Kelly et al., 2021; Montalti et al., 2021; A. M. Scherer et al., 2021; Teasdale et al., 2021; Yigit et al., 2021). Also, parents from Black, Asian or minority ethnic groups were more hesitant to vaccinate their children against COVID-19 (S. Bell et al., 2020; Kelly et al., 2021; A. M. Scherer et al., 2021).

Positive attitudes with regards to vaccination affected positively parents’ intention to vaccinate their children against COVID-19. In particular, children’s complete vaccination history, recent history of vaccination against influenza, confidence in vaccines, confidence in COVID-19 vaccine safety and efficacy, and COVID-19 vaccine uptake among parents were associated with increased intended uptake of a COVID-19 vaccine (Goldman et al., 2020; Hetherington et al., 2021; Kelly et al., 2021; Ruggiero et al., 2021; Skjefte et al., 2021; Szilagyi et al., 2021; Yilmaz & Sahin, 2021; Zhang et al., 2020). On the other hand, overall vaccination hesitancy, and concerns for serious side effects and effectiveness of COVID-19 vaccination decreased parents’ willingness to vaccinate their children against COVID-19 (Montalti et al., 2021; Ruggiero et al., 2021; Zhang et al., 2020).

Higher levels of perceived threat from the COVID-19, worry, fear, and anxiety were associated with parents’ intention to accept COVID-19 vaccination for their children (Kelly et al., 2021; Skjefte et al., 2021; Yigit et al., 2021; Yilmaz & Sahin, 2021). Also, parents with higher levels of knowledge about prevention measures, information about the COVID-19 pandemic, confidence in public health agencies/health science, and compliance to mask guidelines were more likely to vaccinate their children (Brandstetter et al., 2021; Skjefte et al., 2021).

## Discussion

To our knowledge, this is the first systematic review and meta-analysis that assesses the willingness of parents to vaccinate their children against the COVID-19 and investigates the predictors for their decision. Seventeen papers including 45,783 parents met our inclusion criteria. The primary reasons that papers were excluded from our systematic review include other types of publications (e.g. reviews, qualitative studies, case reports, protocols, etc.) and irrelevant research question.

We found that the overall proportion of parents that intend to vaccinate their children against the COVID-19 is moderate (56.8%) with a wide range among studies from 29% to 72.7%. Parents’ intention to vaccinate their children against the COVID-19 is lower than intention of the general population to take a COVID-19 vaccine (56.8% vs. 73%) (Snehota et al., 2021; Wang, Yang, et al., 2021). Also, the willingness of high-risk groups such as healthcare workers to accept COVID-19 vaccination is higher than parents’ willingness to vaccinate their children (63.5% vs. 56.8%) (Galanis et al., 2020; Luo et al., 2021). A possible explanation for the lower overall intention of parents to vaccinate their children against the COVID-19 demonstrated by our meta-analysis could be the perception of a very low risk of severe COVID-19 in children and the fact that children are often asymptomatic carriers. The wide range of parents’ willingness among studies is confirmed by similar reviews in the general population and could be due to different study designs, study populations, levels of knowledge and information, attitudes towards vaccination etc. (Galanis et al., 2021; Snehota et al., 2021; Wang, Yang, et al., 2021). Interestingly, we did not find differences in parents’ willingness to vaccinate their children according to the continent that studies were conducted. However, the low number of studies including in our subgroup analysis could affect this finding and thus more studies should be performed to infer more valid conclusions.

It is noteworthy that our meta-regression analysis revealed that data collection time does not affect parents’ intention to vaccinate their children but studies of current and ongoing attitudes towards COVID-19 vaccination should be conducted since information and knowledge about COVID-19 vaccines are still evolving.

According to our review, several socio-demographic characteristics affect parents’ willingness to vaccinate their children against COVID-19. In particular, mothers and older parents were more hesitant, a finding that is confirmed by the literature since females and older individuals are in general more likely to report vaccine hesitancy (Galanis et al., 2021; C. Lin et al., 2020; Neumann-Böhme et al., 2020; Schwarzinger et al., 2021). This could be due to the fact that males and older individuals, reported to be at higher risk of intensive care unit admission and death from COVID-19, and so could be more prone to vaccination (Bienvenu et al., 2020; Peckham et al., 2020). On the other hand, females tend to experience more adverse events after COVID-19 vaccination and their vaccine hesitancy may be related with poor knowledge regarding issues such as fertility, pregnancy, and breastfeeding (Schrading et al., 2021; B. Xu et al., 2021). Therefore, mothers could be more worried about potential side effects of the COVID-19 vaccines in their children, and thus are more reluctant to vaccinate their children.

Moreover, we found that higher educational level is associated with increased intended uptake of a COVID-19 vaccine. Impact of parents’ educational level on vaccine hesitancy is a controversial issue since previous studies have shown that lower educational level is associated with more concerns about vaccine safety and efficacy (Gust et al., 2003; Shui et al., 2006; Smith et al., 2004), but other studies found the opposite (Opel et al., 2011). Also, a higher level of parents’ education is related with higher confidence toward vaccination by giving more tools for decision-making (Bocquier et al., 2018; Gualano et al., 2018; Kempe et al., 2020), but higher educated parents are more likely to forego immunizations (Gilkey et al., 2013; Smith et al., 2011).

Our review revealed that parents from Black, Asian and minority ethnic groups are less willing than White parents to vaccinate their children against COVID-19. This is consistent with a systematic review which shows that COVID-19 vaccination uptake is higher among individuals from White race than individuals from Black race (Galanis et al., 2021). Also, individuals from Black, Asian and minority ethnic groups have a lower level of COVID-19 vaccine acceptability (Funk & Tyson, 2021; Hamel et al., 2021; Malik et al., 2020; Ruiz & Bell, 2021) and they have lower seasonal influence vaccine coverage (Williams et al., 2017). Given that people from Black, Asian and minority ethnic groups are at higher risk of acquiring SARS-CoV-2 infection and at increased risk of adverse outcomes from COVID-19, a concerted effort must be made to minimize inequalities in COVID-19 vaccination uptake and ensure equitable access to the COVID-19 vaccines (Martin et al., 2020; Sze et al., 2020; Voysey et al., 2021).

We found that parents’ positive attitudes towards vaccination affect their decision to vaccinate their children against COVID-19. In particular, parents whose children had recently received the influenza vaccination or had a completed vaccination history reported a higher likelihood of COVID-19 vaccination for their children. During the COVID-19 pandemic, an important predictor of future behavior remains past behavior (Bourassa et al., 2020). Past behavior predicts future behavior in a direct pathway, where a habitual process occurring, or in an indirect pathway via conscious, intentional processes (Ouellette & Wood, 1998; Schwarzer & Hamilton, 2020). For instance, several studies have identified the relation between individuals’ vaccination in the past and uptake of the pandemic H1N1 vaccine (Bish et al., 2011; Rubin et al., 2011; Setbon & Raude, 2010; Torun et al., 2010). This pattern is similar to our finding that COVID-19 vaccine uptake among parents is associated with increased intended uptake of a COVID-19 vaccine among children. Moreover, the COVID-19 pandemic seems to increase polarization of parents’ vaccination behaviors since parents who did not vaccinate their children in the past reported becoming even less likely to vaccinate them in the near future (Sokol & Grummon, 2020).

According to our review, confidence in vaccination, concerns for serious side effects and effectiveness of vaccines, and vaccine hesitancy are significant predictors of parents’ attitudes regarding vaccination. These findings are confirmed by the literature since parents in the USA are hesitant to vaccinate their children with routine immunizations because of safety, side effects and low effectiveness concerns (Kempe et al., 2020; Nyhan & Reifler, 2015). Vaccine hesitancy is a complex issue and one of the main obstacles to control the COVID-19 pandemic since an instrumental percentage of the general population refuses COVID-19 vaccines (Jaca et al., 2021; Wiysonge et al., 2021). Unfortunately, providing information on vaccine safety and effectiveness to individuals who are vaccine-hesitant can be counterproductive (Nyhan et al., 2014; Nyhan & Reifler, 2015; L. D. Scherer et al., 2016). Tailored and targeted communication materials, and balanced information on vaccines providing both the benefits and risks of vaccination are necessary to optimize vaccine uptake (Dubé et al., 2015, 2020). A robust, transparent, reasonable, and widespread COVID-19 vaccine educational campaign harnessing media, healthcare workers, leaders, and social influencers should be implemented by the public health officials to diminish parents’ concerns for COVID-19 vaccine safety and efficacy (Schaffer DeRoo et al., 2020). Also, behavioral-change theories (e.g., the health-belief model) have already been effectively adapted to improve individual medical use and should be used by government and health authorities to curb COVID-19 vaccine hesitancy among parents (L. Lin et al., 2020; Opel et al., 2009).

Since COVID-19 vaccine safety and effectiveness are key parental concerns, it is critical to emphasize the safety profile of COVID-19 vaccines for children based on evidence from randomized controlled trials and post-approval data. Well-informed parents experience less worry, fear, and anxiety about COVID-19 and are more likely to receive a COVID-19 vaccine for their children as suggested by our review. The rigorous development and approval process of COVID-19 vaccines by the federal agencies worldwide increase parents’ concerns and there is a need for continued transparency and active public education regarding the COVID-19 vaccines development (B. P. Bell et al., 2020; Lee et al., 2020). In that case, the role of primary care physicians to communicate about COVID-19 vaccines for children is critical since prior studies show that clear messages and recommendations by primary care physicians have a large impact on vaccine uptake (Braun & O’Leary, 2020; Dempsey & O’Leary, 2018; Edwards et al., 2016).

## Limitations

This systematic review has several limitations. In particular, findings of our review could not be generalized since the number of studies is relatively low and studies were conducted mainly in the USA and China. Moreover, the statistical heterogeneity was very high due probably to heterogeneity in study designs and populations. To account for this heterogeneity, we applied a random effects model and we performed subgroup and meta-regression analysis. Unfortunately, due to the limited number of studies, we performed subgroup analysis only for two variables. At least, subgroup analysis and leave-one-out sensitivity analysis revealed that our results are robust. We searched for studies conducted till to August 11, 2021 but availability of COVID-19 vaccines and evidence from randomized controlled trials and post-approval data are increasing on an on-going basis and parents’ attitudes could be changed. Thus, our findings may not be generalizable to later in the COVID-19 pandemic. Since all studies in our review were cross-sectional, we cannot infer causal relationships between parents’ willingness to vaccinate their children against the COVID-19 and predictors of this attitude. We consider predictors of parents’ intention to vaccinate their children as a potential area for future study since only socio-demographic variables have so far been investigated thoroughly. Future studies should assess broader and diverse parent populations to fully understand the factors that affect parents’ intention to vaccinate their children against the COVID-19. Finally, the proportion of parents that agreed to vaccinate their children against the COVID-19 may be a biased estimation since studies measured willingness and not COVID-19 vaccination uptake.

## Conclusions

High vaccination coverage is indispensable to control the COVID-19 pandemic. Given the highly transmissible delta variant, COVID-19 vaccination coverage should be increased to achieve herd immunity to COVID-19. This is the main reason that the COVID-19 vaccine rollout is just expanding to the children population. Thus, it is critical to better understand what factors affect parents’ decision to vaccinate their children against COVID-19. Understanding parental COVID-19 vaccine hesitancy does help policy makers to change the stereotypes and establish broad community COVID-19 vaccination. As global COVID-19 vaccines rollout continue, our review could help policy makers and healthcare workers to understand parental decision around COVID-19 vaccination. This information can be used for evidence-based targeted campaigns and health interventions to ultimately maximize future COVID-19 vaccine uptake among children. There is a need to build vaccine confidence during the COVID-19 pandemic through clear messages and effective community engagement. Targeted public health strategies should aim to assuage parents’ concerns regarding COVID-19 vaccines. Identification of the factors that affect parents’ willingness to vaccinate their children against COVID-19 will provide opportunities to enhance parents trust in the COVID-19 vaccines and optimize children’s uptake of a COVID-19 vaccine.

## Data Availability

Data will be available after a reasonable request.

---

**Supplementary Figure S1.**
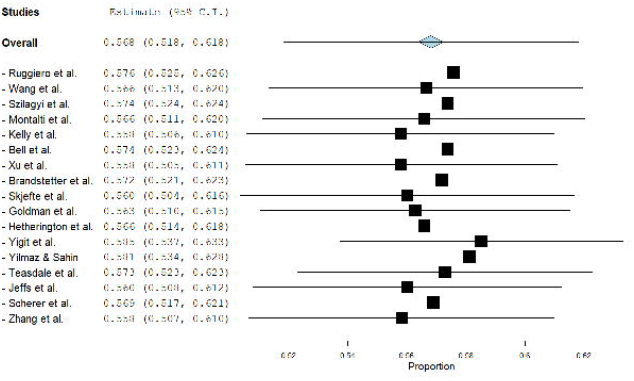
A leave-one-out sensitivity analysis of parents’ willingness to vaccinate their children against the COVID-19.

**Supplementary Figure S2.**
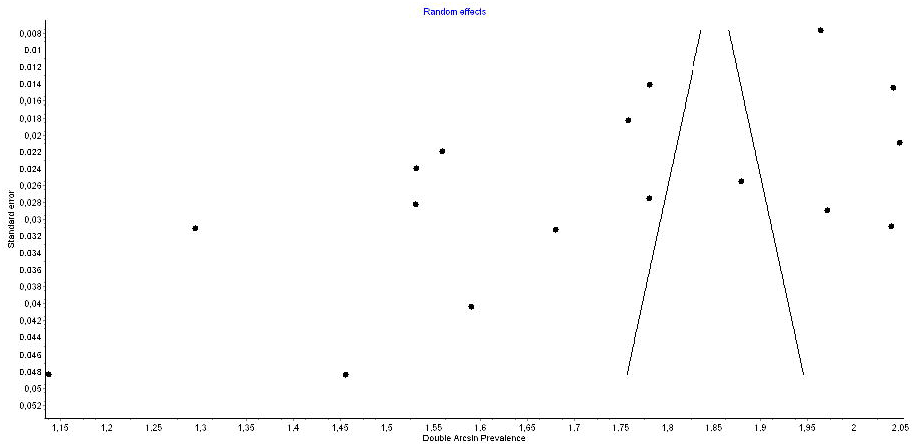
Funnel plot of parents’ willingness to vaccinate their children against the COVID-19.

**Supplementary Table 1.**
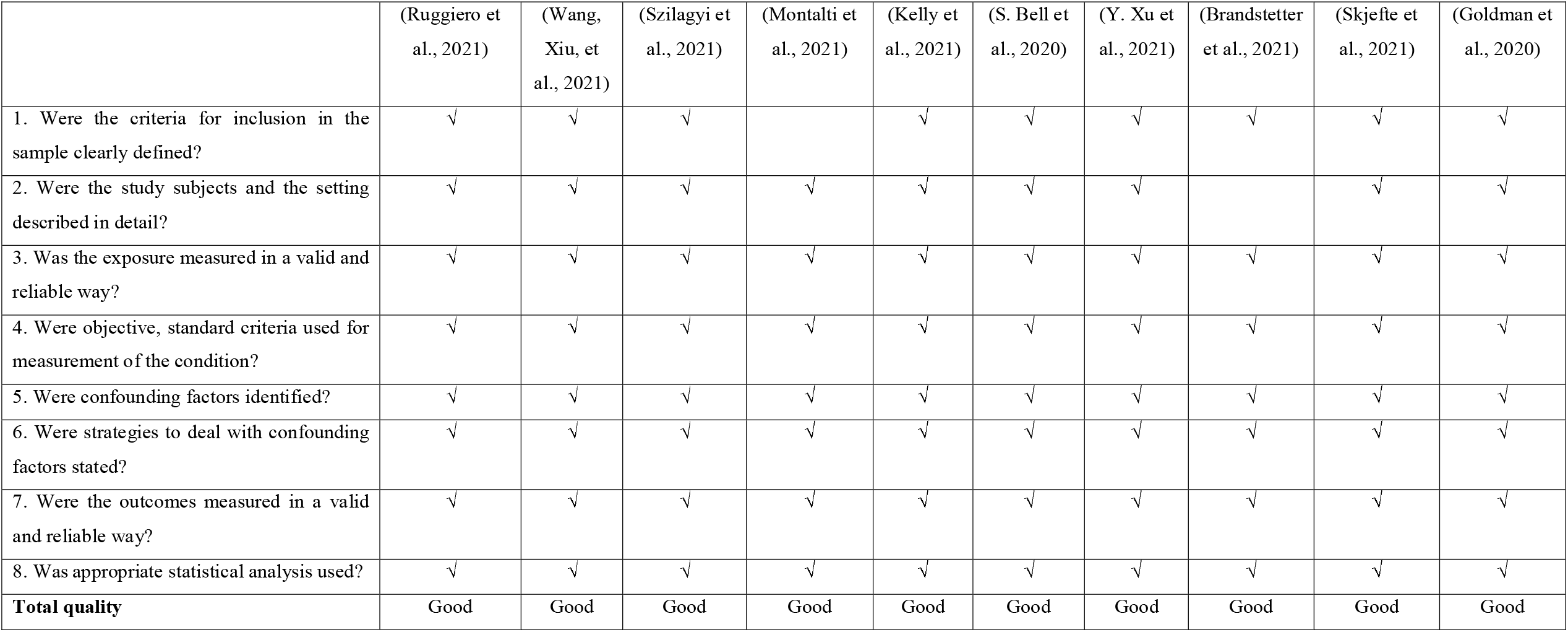
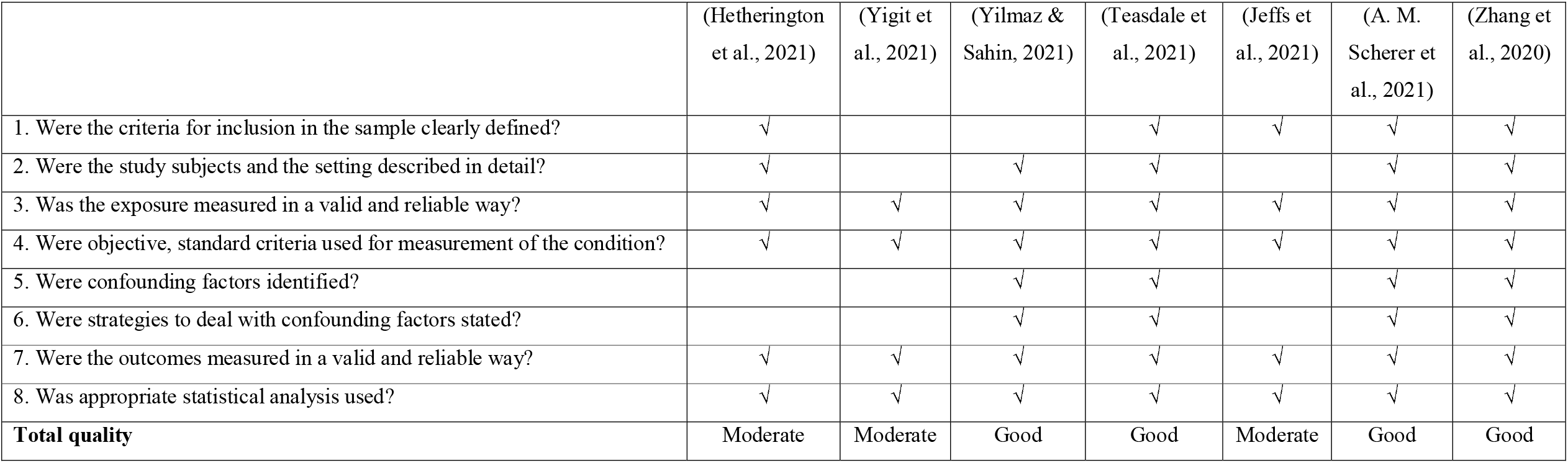
Quality of studies included in this systematic review.

